# COVID-19: the key role of pulmonary capillary leakage. An observational cohort study

**DOI:** 10.1101/2020.05.17.20104877

**Authors:** Maddalena Alessandra Wu, Tommaso Fossali, Laura Pandolfi, Luca Carsana, Davide Ottolina, Vanessa Frangipane, Roberto Rech, Antonella Tosoni, Andrea Agarossi, Chiara Cogliati, Federica Meloni, Beatrice Marchini, Manuela Nebuloni, Emanuele Catena, Riccardo Colombo

## Abstract

**Background:** COVID-19 induces progressive hypoxemic respiratory failure and acute respiratory distress syndrome, mostly due to a dysregulated inflammatory response. Since the first observations of COVID-19 patients, significant hypoalbuminemia was detected. This study aimed to investigate the hypothesis that hypoalbuminemia in COVID-19 patients is due to pulmonary capillary leakage and to test its correlation with indicators of respiratory function.

**Methods:** 174 COVID-19 patients, 92 admitted to the Intermediate Medicine ward (IMW), and 82 to the Intensive Care Unit (ICU) at Luigi Sacco Hospital in Milan were included in this study.

**Findings:** Serum albumin concentration was decreased in the whole cohort, with ICU patients displaying lower values than IMW patients [20 (18-23) vs 28 (24-33) g·|^-1^, p<0.001], Lower albumin values were found in patients belonging to a more compromised group (lower PaO_2_ to FiO_2_ ratio and worst chest X-ray findings). In a subset of 26 patients, analysis of bronchoalveolar lavage fluid (BALF) highlighted high protein concentrations, which were correlated to Interleukin-8 and Interleukin-10 BALF concentration. The length of hospitalisation [20 (15-29) vs 8 (5-14) days, p<0.0001] and death rate (52.4% vs 21.7%, p<0.0001) were higher in ICU than in IMW patients, while a strict relation between hypoalbuminemia and 30 day-survival was detected in the whole cohort. Electron microscopy examinations of eight out of ten autopsy lung tissues showed diffuse loosening of interendothelial junctional complex.

**Interpretation:** The degree of hypoalbuminemia can be considered as a useful severity marker in hospitalised COVID-19 patients. Pulmonary capillary leak syndrome secondary to the hyperinflammatory state plays a key role in the pathogenesis of COVID-19 respiratory dysfunction and should be regarded as a therapeutic target.

## INTRODUCTION

The outbreak of the new Severe Acute Respiratory Syndrome Coronavirus 2 (SARS-CoV-2) infection begun in December 2019 in Wuhan, China (1), and rapidly spread to every continent, leading the scientific community to conduct multidisciplinary studies to investigate its pathogenesis and address issues related to the most appropriate management.

SARS-CoV-2 disease (COVID-19) usually progresses through subsequent phases (early infection with viral response phase, pulmonary phase, and hyperinflammation phase (2). In its most severe presentation, COVID-19 induces progressive hypoxemic respiratory failure ranging from mild pneumonia to acute respiratory distress syndrome (ARDS), mostly due to a dysregulated inflammatory response (3, 4). During the hyperinflammation stage, many proinflammatory cytokines, chemokines, and mediators (such as interleukin 6, interleukin 1β, tumour necrosis factor (TNF)-α, granulocyte-colony stimulating factor, macrophage inflammatory proteins 1-α) have been shown to be significantly increased in peripheral blood, likely playing a pivotal role not only in respiratory injury but also in multiple organ failure. These molecules cause remarkable damage especially to those sites with high expression of angiotensin-converting enzyme 2 (ACE2), the functional receptor for SARS-CoV-2 cell entry, leading to the clinical picture of the so-called “viral sepsis” (5, 6).

Thanks to autopsy studies, direct viral damage was also confirmed by the identification of viral particles in the bronchial and type 2 alveolar epithelial cells by electron microscopy (7).

Recently, interest has been raised on the role of endothelial dysfunction in COVID-19. The dysfunction appears to be the result of multiple concomitant mechanisms, including reaction to the invading pathogen as well as to hypoxia, activation and recruitment of immune cells (mononuclear cells and neutrophils), with the production of inflammatory mediators (including cytokines, polyphosphates, neutrophil extracellular traps –NETS) (8), and damage to cells releasing histones (9). These hits confer to the vascular endothelium a procoagulant vasoconstriction-prone inflammatory status (10, 11). The involvement of endothelial cells across vascular beds of different organs in a series of COVID-19 patients has been shown, with the presence of viral elements within endothelial cells inducing cell death of both endothelial and immune cells (apoptosis, NETosis) (12). This process may lead to the loss of integrity of the epithelial-endothelial (air-blood) barrier, with exudate in the alveolar cavity, a mechanism which is consistent with imaging findings, showing signs of interstitial-alveolar damage (B lines, white lung and patchy pattern at lung ultrasound, ground-glass opacities and hazy areas with slightly increased density at CT scan) (13, 14). The nature of the above-mentioned alveolar exudate has been a matter of investigation.

Since our very first observations of COVID-19 patients in the last days of February 2020, we found significant hypoalbuminemia, both in patients admitted to Intermediate Care Wards (IMW, Internal and Emergency Medicine Departments) and in those rapidly admitted to Intensive Care Units (ICUs). No proteinuria or protein-losing enteropathy of such an entity to justify so significant hypoalbuminemia was ever detected. Remarkably, the administration of albumin did not seem to be able to restore intravascular albumin concentration.

We conducted this multidisciplinary study to investigate the hypothesis that COVID-19 is able to induce pulmonary capillary leakage which, despite being secondary to the hyperinflammatory state and the cytokine storm, significantly contributes to the pathogenesis of COVID-19-related clinical as well as laboratory and imaging pictures.

## MATERIALS AND METHODS

### Study design and participants

This is a retrospective, observational cohort study carried out at Luigi Sacco Hospital, a referral centre for highly transmissible diseases in Milan, Italy, during the large COVID-19 epidemic surge in Northern Italy. Hospital charts of all patients ≥18 years of age admitted to the Intensive Care Unit (ICU) and Intermediate Medical Ward (IMW) between February 21 and April 15, 2020, were screened. Inclusion criteria were SARS-CoV-2-induced lung disease confirmed by real-time PCR on throat swab samples, and a serum albumin measurement within 72 hours from hospital admission. Exclusion criteria were age < 18 years and missing serum albumin measurements in the first 72 hours since hospital admission. We compared the clinical characteristics of ICU and IMW patients.

Research and data collection protocols were approved by the Institutional Review Board (Comitato Etico di Area 1). Written informed consent was obtained by survivors and waived in all others.

For autopsies, the study followed the Italian general rules used for research related to scientific purposes (official regulations n.72-26/03/2012).

### Procedures

Diagnostic and therapeutic interventions were chosen by the treating physician according to current clinical practice. Patients were admitted to ICU if they failed a trial with continuous positive airway pressure by a helmet. Failure was defined as respiratory rate >30 breaths per minute and PaO_2_ to FiO_2_ ratio <150, or respiratory acidosis with pH<7.36 and PaCO_2_ >50 mmHg, or agitation, or confusion.

Demographic features, laboratory results on admission, need for organ support, as well as ICU and hospital length of stay (LOS), and outcomes were retrieved from hospital charts.

Chest X-rays (CXR) performed at hospital admission were independently reviewed by two physicians with experience in respiratory medicine, and a CXR scoring system specifically designed for semi-quantitative assessment of lung disease in COVID-19 (Brixia score) was calculated. This ranks the pulmonary involvement on an 18-point severity scale according to the extent, and characteristics of lung infiltrates (15). Bronchoalveolar lavage was collected from mechanically ventilated ICU patients according to the clinical requirement through disposable bronchoscope aScope™ 4 (Ambu A/S, Baltorpbakken, Denmark). Samples were centrifuged at 400 g for 10 min at room temperature.

To quantify interleukin 8 (IL-8) and interleukin 10 (IL-10), we used SimpleStep ELISA® kit (Abcam, Cambridge, UK). Briefly, 50 μL of each sample was added into ELISA kit wells also adding 50 μL antibody cocktail. After 1 hour at room temperature on a plate shaker, three washes were done to eliminate the unbounded antibody. 100 μL of the substrate was incubated for 10 min in the dark at room temperature on a plate shaker, followed by 100 μL stop solution to read the absorbance at 450 nm. For IL-8 analyses, samples were diluted (from 1:10 to 1:1000).

To quantify proteins in BALF samples, Pierce™ BCA protein assay kit (Thermo Fisher Scientific, Massachusetts – USA) was used. The standard range used was from 20 to 2000 μg·ml^-1^. 25 μl of each sample was added to a 96-well plate and 200 μl of BCA working reagent. After 30 min at 37°C, absorbance was read at 550 nm.

We collected lung tissues of ten patients who died in ICU. Tissues were fixed in 10% buffered formalin and embedded in paraffin. Three-μm paraffin sections were stained by hematoxylin-eosin for histological examination. Additional samples were fixed in 2.5% glutaraldehyde and prepared for ultrastructural analysis. Thin sections were stained with uranyl acetate/lead citrate and examined by EM-109 ZEISS and CCD-Megaview G2 (I-TEM imaging platform software).

### Statistical analysis

Descriptive statistics were used to characterise the cohort of patients, with denominators reflecting the number of patients treated either in ICU or IMW. Categorical data were compared using Fisher’s exact test or a χ^2^ test. Between-group comparison of continuous variables was accomplished via Mann-Whitney test or one-way analysis of variance, as appropriate. Correlation between non-normally distributed variables was assessed with Spearman’s Rho. Comparison of survival curves was analysed with the Mantel-Cox test. Values are shown as median and (interquartile range). Statistical significance was defined as p<0.05. Data were analysed with Graphpad Prism version 8.4.1.

## RESULTS

From February 21 to April 15, 2020, 174 hospitalised patients with COVID-19 related respiratory symptoms met the inclusion criteria. Of these, 92 were admitted to the IMW and 82 to ICU. All patients came from the area of a large epidemic outbreak located in Lombardy, Northern Italy. Characteristics of the studied population at admission are shown in Table 1. Patients’ age did not differ between the ICU and the IMW population. There was a marked prevalence of male gender among ICU patients, while there was a higher prevalence of a history of smoking, cardiovascular diseases, and diabetes among IMW patients. The time elapsed from the onset of the respiratory symptoms to hospital admission was shorter in ICU compared to IMW patients [6 (4-9.2) vs 8 (6-11) days, p=0.019], while no significant difference was detected in the time from hospital admission to starting continuous positive airway pressure treatment [1 (0-3) vs 1 (0-2) days, p=0.99]. Positive end-expiratory pressure at the beginning of ventilatory support was higher in ICU than IMW patients [13.5 (12-16) vs 12 (10-12.5) cmH_2_O, p<0.0001], D-dimer, lactate dehydrogenase, aspartate aminotransferase, and C-reactive protein were higher in ICU patients, as well as white blood cell and neutrophil count. However, in ICU patients lymphopenia was more remarkable than in IMW patients [661 (460-935) vs 1062 (760-1513) lymphocytes·ml^-1^, p<0.0001]. None of the patients met the diagnostic criteria of the International Society on Thrombosis and Haemostasis for disseminated intravascular coagulation. Median serum albumin concentration was below the laboratory reference value (30 g·|^-1^) in the whole cohort, but ICU patients displayed more pronounced hypoalbuminemia than IMW patients [20 (18-23) vs 28 (24-33) g·|^-1^, p<0.0001]. The degree of respiratory involvement, expressed by the partial pressure of arterial oxygen (PaO_2_) to the fraction of inspired oxygen (FiO_2_) ratio, and the Brixia score, was worst in ICU patients.

**Table 1.** Characteristics of the studied population.

|  | ICU (n=82) | IMW (n=92) | p |
| --- | --- | --- | --- |
| Age, years | 62.5 (48.7-68) | 60 (49-73) | 0.68 |
| F/M | 17 (20.7)/65 (79.3) | 36 (39.1)/56 (60.9) | 0.008 |
| BMI | 27.8 (25.4-31.9) | 26.6 (23.9-29.8) | 0.35 |
| **Comorbidities, n (%)** |  |  |  |
| Smokers | 11 (13.4) | 47 (51.1) | <0.0001 |
| Cardiovascular disease | 39 (47.6) | 41 (44.6) | 0.76 |
| Respiratory disease | 3 (3.7) | 12 (13) | 0.03 |
| Diabetes | 11 (13.4) | 26 (28.3) | 0.025 |
| Cancer | 5 (6.1) | 12 (13) | 0.13 |
| HIV | 2 (2.4) | 4 (4.3) | 0.68 |
| Immunosuppressive therapy | 4 (4.9) | 4 (4.3) | 1 |
| Symptoms to hospital admission, days | 6 (4-9.2) | 8 (6-11) | 0.019 |
| Hospital admission to CPAP, days | 1 (0-3) | 1 (0-2) | 0.99 |
| CPAP to mechanical ventilation, days | 2 (1-4) | 3 (1.5-4) | 0.73 |
| Mechanical ventilation, n (%) | 78 (95.1) | 4 (4.3) | <0.0001 |
| PEEP, cmH<sub>2</sub>O | 13.5 (12-16) | 12 (10-12.5) | <0.0001 |
| Tidal Volume, ml | 550 (500-600) |  |  |
| Respiratory Rate, bpm | 20 (18-24) | 20 (18-26) | 0.1 |
| Respiratory System Compliance, ml·cmH<sub>2</sub>O<sup>-1</sup> | 38 (31-46) |  |  |
| Fibrinogen, mg·dl<sup>-1</sup> | 700 (700-700) | 700 (600-700) | 0.21 |
| D-dimer, ng·ml<sup>-1</sup> | 2374 (1186-7097) | 950 (490-2320) | <0.0001 |
| Haemoglobin, g·dl<sup>-1</sup> | 12.7 (11.5-13.8) | 12.9 (12.2-14.08) | 0.93 |
| Hematocrit, % | 38 (34-41) | 38.5 (36-41) | 0.12 |
| Platelets, 10<sup>3</sup> cells·ml<sup>-1</sup> | 230 (173-309.7) | 205 (137.2-303.2) | 0.11 |
| WBC, cells·ml<sup>-1</sup> | 8375 (6058-12013) | 5885 (4500-9505) | <0.0001 |
| Neutrophils, cells·ml<sup>-1</sup> | 7526 (4922-10547) | 3910 (2440-7185) | <0.0001 |
| Lymphocytes, cells·ml<sup>-1</sup> | 661 (460-935) | 1062 (760-1513) | <0.0001 |
| S-creatinine, mg·dl<sup>-1</sup> | 0.9 (0.74-1.21) | 0.88 (0.69-1.07) | 0.37 |
| LDH, U·l<sup>-1</sup> | 537 (416-666) | 303 (242-389) | <0.0001 |
| S-Albumin | 20 (18-23) | 28 (24-33) | <0.0001 |
| AST, U·l<sup>-1</sup> | 53 (36-76) | 39 (28-61) | 0.01 |
| ALT, U·l<sup>-1</sup> | 40 (22-72) | 30 (21.75-61.25) | 0.07 |
| S-bilirubin, mg·dl<sup>-1</sup> | 1.2 (1.2-1.2) | 1.19 (1-1.2) | 0.02 |
| S-lactate, mmol·l<sup>-1</sup> | 1.3 (1-1.5) | 1.2 (1-1.5) | 0.63 |
| CRP, mg·l<sup>-1</sup> | 176 (94-278) | 70 (24-140) | <0.0001 |
| arterial-pH | 7.35 (7.29-7.43) | 7.46 (7.43-7.49) | <0.0001 |
| arterial-pO<sub>2</sub>, mmHg | 83 (71-102) | 89.5 (72.25-111) | 0.27 |
| arterial-pCO<sub>2</sub>, mmHg | 46 (40-52) | 38 (34-41) | <0.0001 |
| PaO<sub>2</sub> to FiO<sub>2</sub> ratio | 111 (91-151) | 243 (148-376) | <0.0001 |
| **Chest X-Ray score, n (%)** |  |  |  |
| 0-5 | 1 (1.2) | 47 (51.1) | <0.0001 |
| 6-11 | 5 (6.1) | 25 (27.2) | <0.0002 |
| 12-18 | 76 (92.7) | 14 (15.2) | <0.0001 |
| missed | 0 | 6 (6.5) | 0.03 |
| New RRT during hospital stay, n (%) | 19 (23.2) | 0 | <0.0001 |
| Tocilizumab, n (%) | 25 (30.5) | 18 (19.6) | 0.11 |
| Corticosteroids, n (%) | 5 (6.1) | 3 (3.3) | 0.48 |
| Remdesivir n (%) | 34 (41.5) | 0 | <0.0001 |
| Hydroxychloroquine, n (%) | 60 (73.2) | 79 (85.9) | 0.057 |
| Lopinavir/ritonavir, n (%) | 53 (64.6) | 43 (46.7) | 0.22 |
| **ICU outcome** |  |  |  |
| Mechanical ventilation length, days | 11 (5-16) |  |  |
| LOS, days | 13 (7.7-17) |  |  |
| Dead, n (%) | 41 (50) |  |  |
| still in ICU, n (%) | 3 (3.7) |  |  |
| **Hospital outcome** |  |  |  |
| LOS, days | 20 (15-29) | 8 (5-14) | <0.0001 |
| Dead, n (%) | 43 (52.4) | 20 (21.7) | <0.0001 |
| still in hospital, n (%) | 17 (20.7) | 0 | <0.0001 |

Data are shown as median (25<sup>th</sup>-75<sup>th</sup> percentiles) or as n (%) when indicated. ICU, intensive care unit; IMW, intermediate medical ward; BMI, body mass index; CPAP, continuous positive airway pressure; PEEP, positive end-expiratory pressure; CRP, C-reactive protein; WBC, white blood cells; CXR, chest X-Ray Brixia score; RRT, renal replacement therapy; LOS, length of stay.

To assess whether patients with worse respiratory impairment presented differences in serum albumin, we divided patients into three groups according to PaO_2_ to FiO_2_ ratio (>250, 250-150, <150) or Brixia score (0-5, 6-11, 12-18): lower albumin values were found in patients belonging to more compromised groups (lower PaO_2_ to FiO_2_ ratio and higher Brixia score) (Figure 1A). Median serum albumin was 31 (27-33) g·|^-1^ in patients with PaO_2_ to FiO_2_ ratio >250, 24 (20.75-27.5) g·l^-1^ in those with PaO_2_ to FiO_2_ ratio between 150 and 250, and 20 (18-24) g·|^-1^ in those with PaO_2_ to FiO_2_ ratio <150. Serum albumin concentration was significantly lower in patients with chest X-ray showing more diffuse and pronounced pulmonary involvement (p<0.0001) (Figure 1B): serum albumin was 31 (26.75-35.25) g·|^-1^ with CXR score <5, 27 (24-32) g·|^-1^ with CXR score between 6 and 11, and 21 (18-24) g·|^-1^ with CXR score ≥12. The hospital LOS was higher in ICU than in IMW [20 (15-29) vs 8 (5-14) days, p<0.0001]. The in-hospital death rate was significantly higher in ICU compared to IMW patients (52.4% vs 21.7%, p<0.0001) After stratification of patients by the median of serum albumin concentration in the whole series (24 g·|^-1^), the survival rate at 30 days from admission was significantly different (53.2% vs 21%, p<0.0001) (Figure 3).

**Figure 1.**
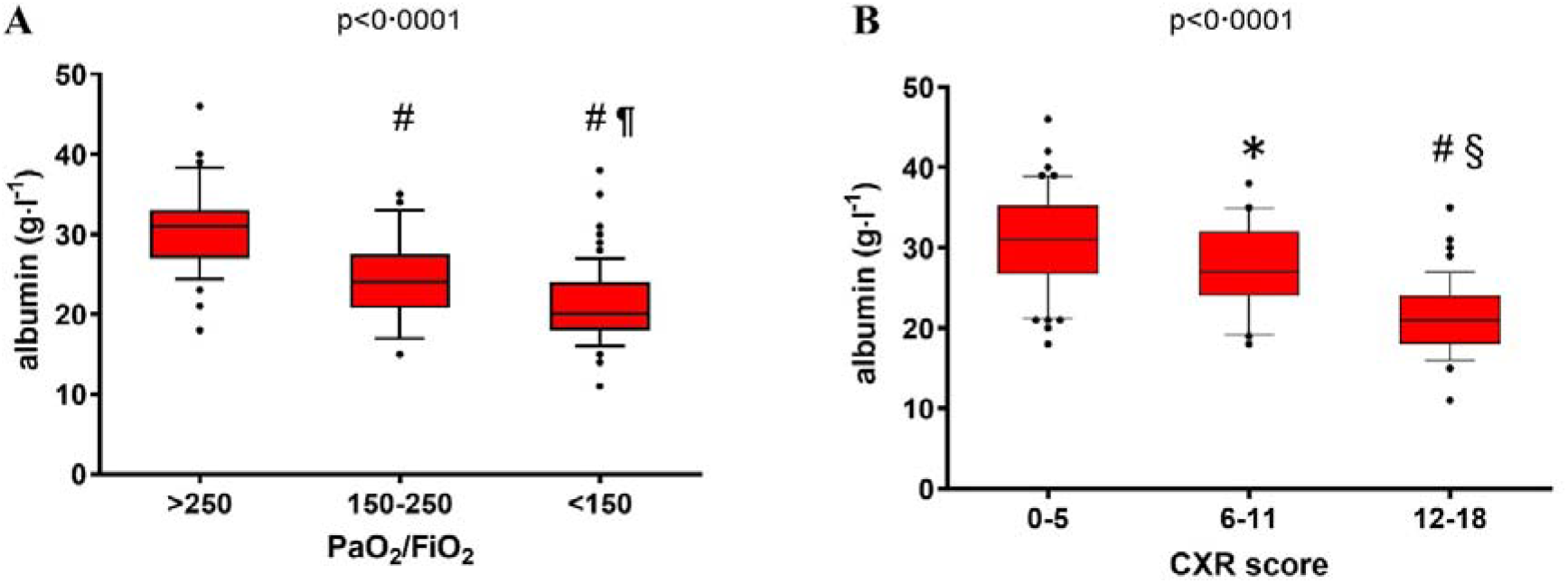
*A)* Serum albumin concentration and PaO_2_ to FiO_2_ ratio (PaO_2_/FiO_2_): serum albumin values were lower in patients with lower PaO_2_/FiO_2_, index of worsening respiratory function (p<0.0001). # p<0.001 vs. PaO_2_ to FiO_2_ >250; ¶ <0.05 vs. PaO_2_ to FiO2 150-250. *B)* Serum albumin concentration and chest X-ray (CXR) score (p<0.0001): higher CXR scores, indexes of more diffuse and clear pulmonary involvement, are found in patients with lower serum albumin concentrations. *p<0.05 vs. 0-5 CXR score; # p<0.001 vs. 6-11 CXR score; § p<0.001 vs. 0-5 CXR score.

In the first 48 hours after admission, qualitative proteinuria measurements were available for 77 ICU patients and were undetectable in 73 (94.8%). Furthermore, quantitative 24-hour proteinuria was available for 12 ICU patients with a median (interquartile range) of 76 (28.5-165) mg.

One patient had a history of chronic enteropathy (Crohn’s disease). No other patients had history or signs of protein-losing enteropathy or malnutrition.

Besides, we measured the concentration of IL-8, IL-10, and proteins in the BALF in 26 critically ill patients during their first week of ICU stay. We found a positive correlation between protein concentration and the proinflammatory IL-8 (ρ=0.61, p=0.001), and an inverse correlation between protein concentration and the anti-inflammatory IL-10 (p=-0.81, p<0.0001) in BALF (Figure 2).

**Figure 2.**
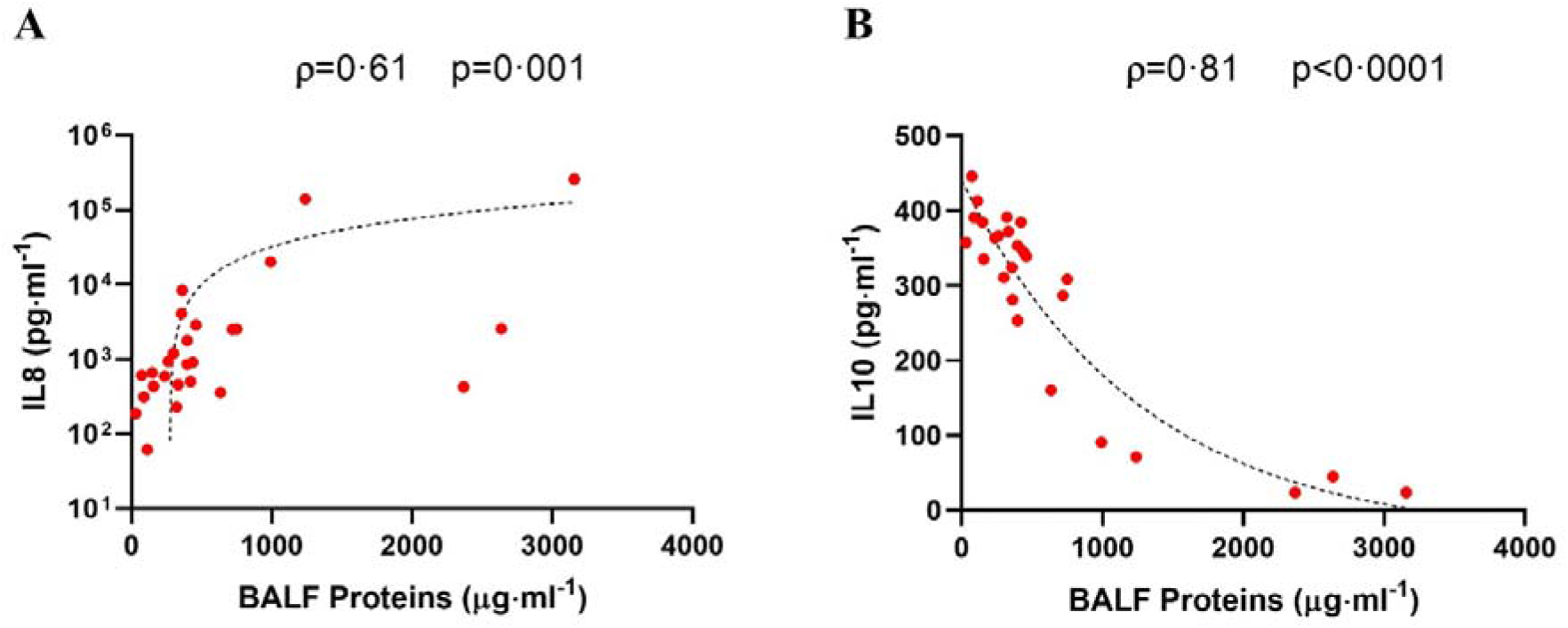
*A)* correlation between concentrations of proinflammatory Interleukin 8 (IL8) and proteins in the bronchoalveolar lavage fluid. *B)* correlation between concentrations of anti-inflammatory Interleukin 10 (IL10) and proteins in the bronchoalveolar lavage fluid (BALF).

**Figure 3.**
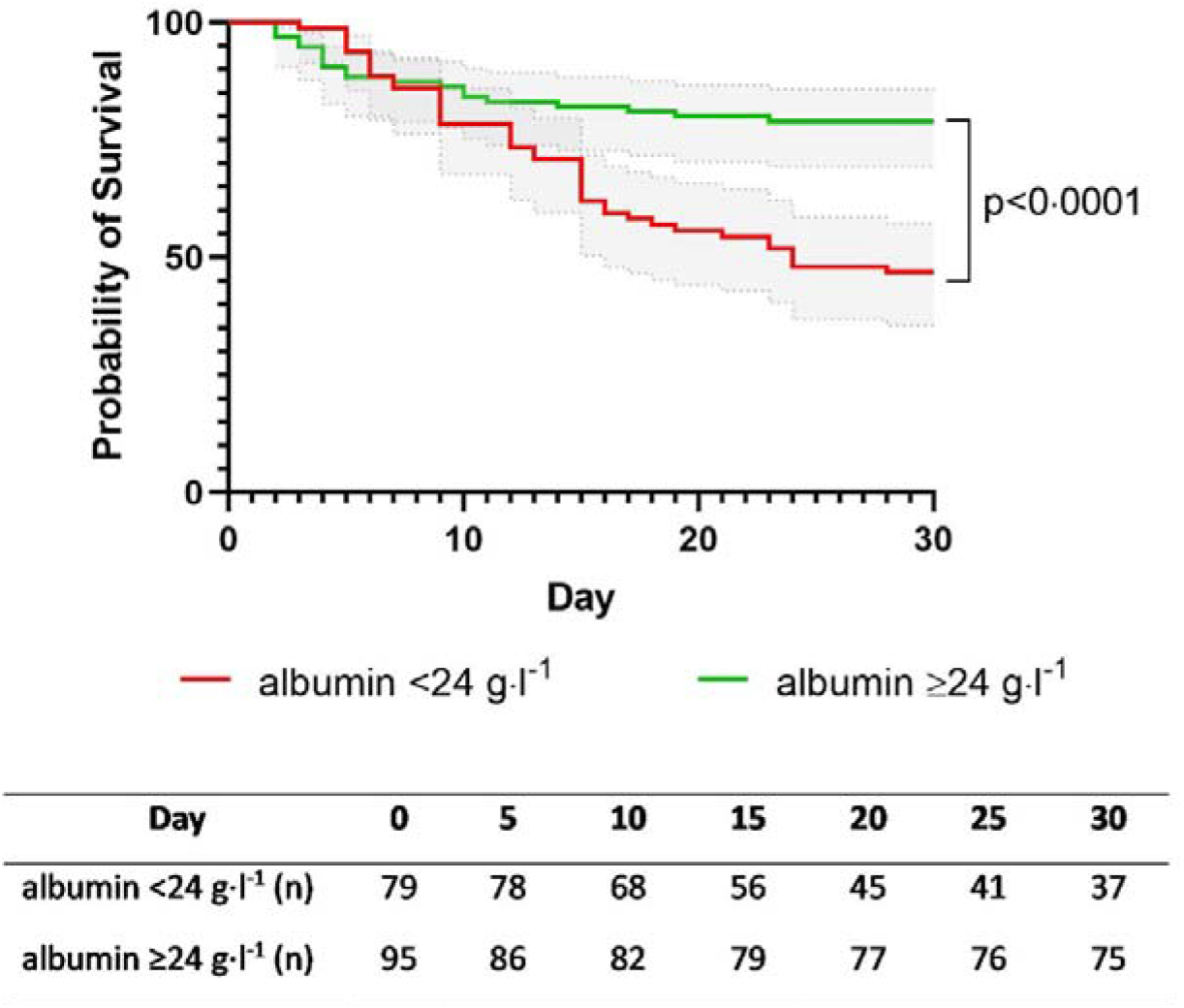
First 30-day survival curves in the whole cohort according to serum albumin concentration at admission. Grey bands represent 95%CI.

Figure 4 (panels A-H) shows the features of ultrastructural and histological samples. At histology, there was proliferation and hyperplasia of type 2 pneumocytes in all the samples; moreover, denudation of alveolar membranes and residual hyaline membrane deposition were observed. The interstitium was focally expanded by mild fibrosis, oedema and moderate lymphocytic infiltrate (Figure 4, panel F).

**Figure 4.**
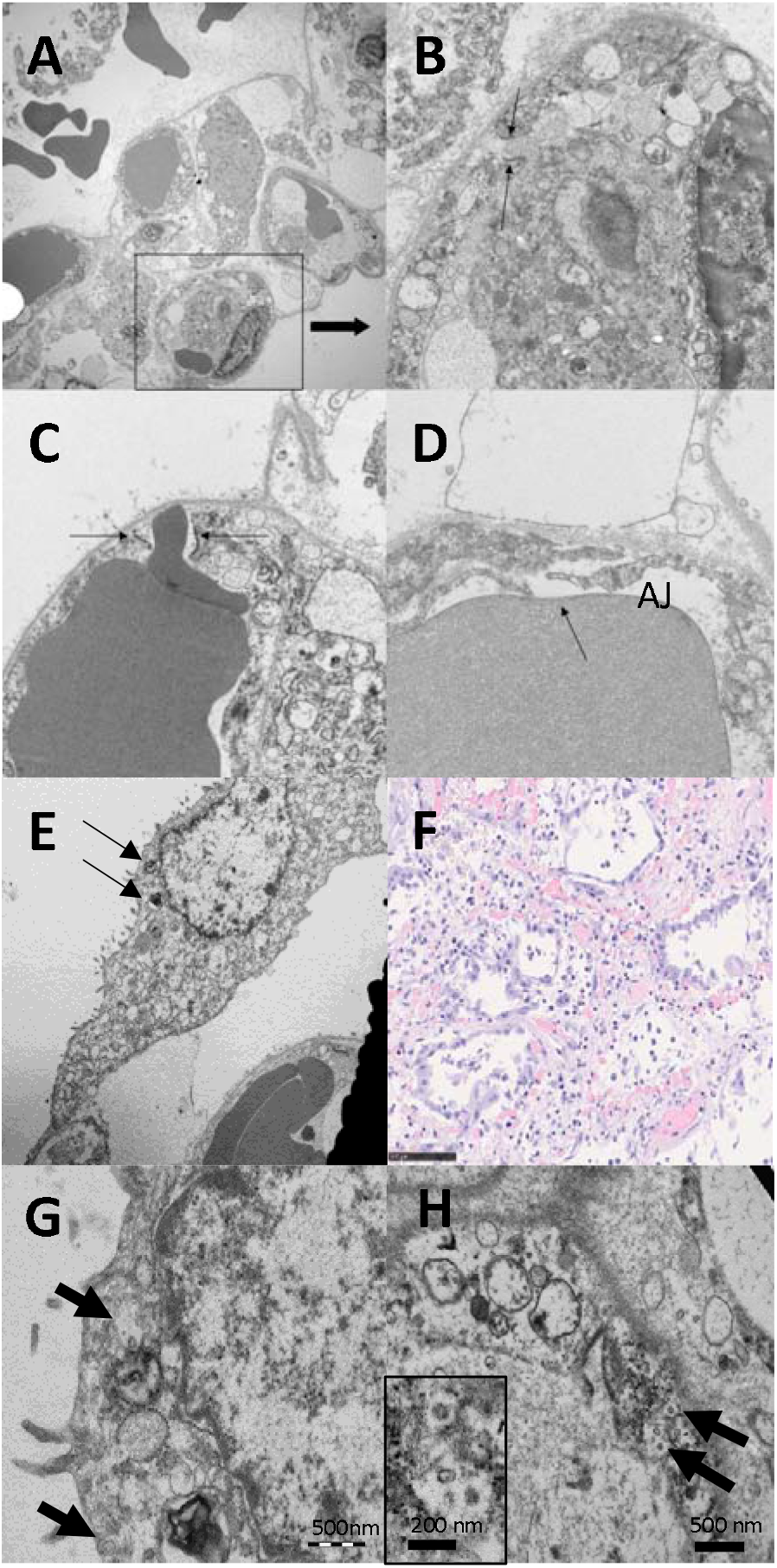
*A-E)* Ultrastructural image of lung septum between two disepithelialised alveoli. One of the capillaries *(A)* shows an evident interruption which at higher magnification *(B)* appears caused by the opening of the junctions. In panels *B* and *C*, the opened junctional complexes still show traces of the cytoskeletal structures (vimentin) of the adherent junctions that appear as an electrondense thickening of the basement membrane. The passage of red cells *(C)* can be observed through the open junction. In panel *D*, there is a small adherent junction still preserved (AJ) while the flaps of the capillary (arrow) are completely separated without a trace of residual junctional structures. *E)* complete detachment of a type 2 pneumocyte from the alveolar basal membrane. Residual small aggregates of surfactant (arrows) are found. *F)* histological picture of lung parenchyma with hyperplastic and atypical type II pneumocytes. Septa are wide for fibrosis ad accumulation of inflammatory cells Panels *G* and *H*: electron microscopy of a pneumocyte *(G)* and an endothelium *(H)* containing numerous virions (arrows) in cytoplasmic vacuoles. Virions had an average diameter of 82 nm, and viral projection about 13nm in length (inset left up, OMx85000). (OM: panel *A* x3000; panels *B-C-D* x12000; panel *E* x4400) panel *G* and *H* x20000 inset panel *H* x50000 Panel *F*: hematoxylin-eosin, OM x20

Electron microscopy of the autopsy lung tissues revealed the extensive opening of junctional complexes (JCs - tight, adherens and gap junctions) in eight out of ten patients (Figure 4) and depletion of surfactant in type 2 pneumocytes of most cases (figure 4, panels A-E). Moreover, viral particles with morphology and intravacuolar localisation coherent with coronavirus (16) were found in the cytoplasm of pneumocytes (panel G) and endothelium (panel H).

## DISCUSSION

The main finding of our study is that hypoalbuminemia frequently found in COVID-19 patients is linked to the degree of respiratory impairment, with a more pronounced decrease in those with a lower PaO_2_ to FiO_2_ ratio or a higher Brixia CXR score. In addition, hypoalbuminemia was associated with higher mortality in IMW; patients admitted to ICU were sicker, had more often hypoalbuminemia and a higher mortality rate as compared to those admitted to the IMW. In fact, the severity of hypoalbuminemia was significantly different according to the degree of respiratory involvement (as assessed by PaO_2_ to FiO_2_ ratio, and imaging patterns).

Hypoalbuminemia is often associated with inflammatory conditions, and it is a common feature of all acutely ill patients. In this scenario, it is linked to both increased vascular permeability (with augmented distribution volume of albumin) and shortened albumin half-life (altered kinetics with neonatal Fc receptor downregulation and increased intracellular breakdown) leading to decreased total albumin mass, despite increased fractional synthesis (17). In hyperinflammatory states, such as trauma, shock or infection, hypoalbuminemia acts as a negative prognostic marker, and hypoalbuminemic patients are more prone to react inadequately to a so-called second-hit, as surgery. Decreased albumin levels are linked to poor outcomes and reduced life expectancy (18). Nonetheless, no clear benefit has been demonstrated for albumin solutions administration in critically ill patients, although a positive effect has been hypothesised in patients with septic shock (19).

A major pathophysiological mechanism of hypoxemia in the acute phase of ARDS due to viral pneumonia (i.e., influenza A) is the direct viral injury to the epithelial–endothelial barrier, resulting in proteinaceous oedema. Usually, the main resistance to protein flux across the epithelial–endothelial barrier is the alveolar epithelium and, to a lesser extent, the endothelial layer, which both depend on the integrity of their JCs (20, 21). However, endothelial cells are the predominant cell line in the lung, accounting for 30% of all cell types; thus, they exert a pivotal role in the regulation of transmembrane flux. Autopsy findings of patients included in this study showed widespread damage of both epithelial and endothelial alveolar cells resulting in a combined injury of both sides of the alveolar-capillary interface. This unfavorable combination allows the passage of fluids and proteins from the intravascular to the alveolar spaces. Moreover, endothelial injury may act as a procoagulant trigger (22), and in association with capillary fluid depletion may ultimately favour intravascular microthrombosis.

Taken together, these results support the hypothesis that, in COVID-19, capillary leak syndrome might significantly contribute to the development of hypoalbuminemia, which in turn could be interpreted as a marker of disease severity.

The term “capillary leak syndrome” refers to leakage of intravascular fluids into the extravascular space. The condition may stem from a variety of underlying clinical conditions and pathophysiological mechanisms: haematological diseases and their complications, drugs and toxic agents (chemotherapy, interleukin 2, Granulocyte-Macrophage Colony-Stimulating Factor, interferon, venom, carbon monoxide), post-surgery and post-trauma states, systemic syndromes (i.e., haemophagocytic lymphohistiocytosis, engraftment syndrome, ovarian hyperstimulation syndrome, differentiation syndrome), dermatologic diseases, infections. In a minority of cases, capillary leakage is considered to be the *primum movens* of the clinical condition, with no identifiable endothelial injury and has transient features: this nosological entity gathers a variety of conditions which are grouped under the denomination of Paroxysmal Permeability Disorders (PPDs) and include primary angioedema, idiopathic systemic capillary leak syndrome (ISCLS), and some rare forms of localised retroperitoneal-mediastinal oedema (23). A variety of clinical pictures may be due to either primary or secondary capillary leak syndrome, ranging from asymptomatic states to generalised oedema and life-threatening multiple organ failure, requiring urgent intervention.

In our COVID-19 cohort, electron microscopy imaging confirms the opening of interendothelial junctions, likely of immune-mediated origin, as corroborated by cytokine detection into the BALFs. These findings are of pivotal importance since it is known that, while fluids and solutes may cross the endothelial barrier via diffusion and filtration, the passage of macromolecules and proteins from the intravascular to the interstitial space is tightly controlled by interendothelial junctions (paracellular pathway) or biochemical-vesicular transport systems (transcellular pathway) (24). Vascular endothelial cadherin (VE-cadherin, also known as CD144) is expressed in essentially all types of vessels and has a crucial role in this highly harmonized system, and its phosphorylation and internalization induced by a variety of triggers underlie alterations of endothelial sieving properties (25). Furthermore, the presence of numerous virions within the cytoplasm of both type 2 pneumocytes and pulmonary endothelial cells in our autopsy series suggests also a direct viral damage to the alveolar-capillary barrier.

High protein concentrations in BALF (with no evidence of proteinuria or protein-losing enteropathy) further support our hypothesis, confirming passage of proteins probably through open JCs.

Some major differences between the suggested pathological mechanism in COVID-19 and what is known about PPDs need to be elucidated. The most important one is related to the site-specificity of COVID-19 endothelial hyperpermeability: the pulmonary vascular bed (as well as cerebral vessels) is usually spared in PPDs, while it is the key player in COVID-19. This might be at least partially due to SARS-CoV-2 tropism, which seems to be closely related to the expression of specific receptors (as ACE2 receptors) on pneumocytes. Nonetheless, the vascular endothelium should be regarded as a complex organ whose phenotypic heterogeneity could also partially explain variable behaviour across different sites (26). Undoubtedly, this different meaningful site-specificity has a substantial impact on the clinical and laboratory picture, since COVID-19 infection only seldom induces overt hypotension and shock, even though signs of hypoperfusion (cold extremities, weak peripheral pulses, metabolic acidosis, prolonged refill time) may be present, and hemoconcentration (which is a key feature of ISCLS) is usually not detected.

Moreover, while ISCLS is considered to be a primarily functional condition, with no detectable damage to endothelial cells (despite reports of elevated levels of proinflammatory mediators and neutrophil granule components in acute ISCLS sera) (27), in COVID-19 a process of programmed cell death and endothelial injury is also involved.

In addition, endothelial injury in COVID-19 patients may underlie diffuse intravascular coagulation of lung microvessels (<1 mm in diameter) (28). Endothelial injury and JCs loosening expose plasma proteins to tissue factor and to the extracellular matrix, which leads to intra-alveolar activation of coagulation and thrombin generation (22), with consequent intravascular coagulation in small vessels, while larger vessels are mainly spared (28).

Our observations pave the way to a more responsible management of COVID-19 patients, avoiding useless (and potentially harmful) administration of albumin (which is likely to extravasate due to loosening interendothelial junctions) and acting on the underlying pathological mechanism. Recently, the Italian Drug Agency approved a study for compassionate use of solnatide, a 17 residue peptide mimicking the lectin-like domain of TNF, which can affect alveolar liquid clearance and reduce the leakage of blood and fluids from the capillaries in the airspace, through activation of the lung epithelial sodium ion channel (ENaC) (29). A phase Mb randomised, placebo-controlled study (EUDRACT No. 2017-003855-47) is already ongoing to investigate the safety and preliminary efficacy of sequential multiple ascending doses of solnatide to treat pulmonary permeability oedema in patients with moderate-to-severe ARDS in Germany and Austria.

Other molecules are under investigation for their ability to stabilise the endothelial barrier. Among those is FX06, a naturally occurring peptide, Bβ15-42, derived from the E1 fragment of fibrin, which binds to VE-cadherin, preventing VE-cadherin-dependent transmigration of leukocytes and stress-induced rearrangement of the endothelial cell actin cytoskeleton leading to rupture of adherens junctions. In experimentally induced ARDS in rodents, lung damage was reduced, and production of inflammatory cytokines (i.e., IL-6) was blunted, while anti-inflammatory cytokine release (i.e., IL-10) was enhanced. Human data from phase I and phase IIa studies evidenced the favourable safety profile of FX06, and this treatment has been successfully applied in a severe case of an Ebola virus-infected patient under compassionate use West Africa and Germany (30).

To our knowledge, this is the first study which, through a multidisciplinary (clinical, radiological, histological) approach, investigates the causes underlying hypoalbuminemia in COVID-19 and addresses its relationship with respiratory impairment. In addition, we are the first authors suggesting a parallel between COVID-19 and PPDs, which, if confirmed, could lead to useful clues for future treatment research. However, we are aware that our study has also some limitations. First, it is retrospective, and our findings will need confirmation in prospective studies. Moreover, we propose a pathophysiological model which necessarily needs validation, i.e. in animal studies.

In conclusion, hypoalbuminemia is a frequent finding in COVID-19 patients and is linked to the severity of lung injury. It might depend on the complex interplay between direct viral effects and the hyperinflammatory host reaction, which lead to endothelial dysfunction and capillary leak syndrome. Further research is needed to get deeper insights into the mechanisms leading to this condition, and to identify new potential therapeutic targets to mitigate or even avoid endothelial dysfunction.

## Data Availability

Data will be available in anonymized form by request to the corresponding author for research purposes, after approval by the institutional ethical committee, and after approval by all co-authors.

## AUTHORS CONTRIBUTION

Concept and design: Colombo and Wu. Acquisition, analysis, or interpretation of data: all authors. Drafting of the manuscript: Wu and Colombo. Critical revision of the manuscript for important intellectual content: all authors. Statistical analysis: Colombo. Supervision: Meloni, Cogliati, Nebuloni, Catena.

The corresponding author had full access to all the data in the study and had final responsibility for the decision to submit for publication.

## REFERENCES

1. Zhu N, Zhang D, Wang W, Li X, Yang B, Song J, et al. A Novel Coronavirus from Patients with Pneumonia in China, 2019. N Engl J Med. 2020;382(8):727–33.

2. Siddiqi HK, Mehra MR. COVID-19 illness in native and immunosuppressed states: A clinical-therapeutic staging proposal. J Heart Lung Transplant. 2020;39(5):405–7.

3. Huang C, Wang Y, Li X, Ren L, Zhao J, Hu Y, et al. Clinical features of patients infected with 2019 novel coronavirus in Wuhan, China. Lancet. 2020;395(10223):497–506.

4. Liu J, Li S, Liu J, Liang B, Wang X, Wang H, et al. Longitudinal characteristics of lymphocyte responses and cytokine profiles in the peripheral blood of SARS-CoV-2 infected patients. EBioMedicine. 2020;55:102763.

5. Hamming I, Timens W, Bulthuis ML, Lely AT, Navis G, van Goor H. Tissue distribution of ACE2 protein, the functional receptor for SARS coronavirus. A first step in understanding SARS pathogenesis. J Pathol. 2004;203(2):631–7.

6. Li H, Liu L, Zhang D, Xu J, Dai H, Tang N, et al. SARS-CoV-2 and viral sepsis: observations and hypotheses. Lancet. 2020.

7. Yao XH, Li TY, He ZC, Ping YF, Liu HW, Yu SC, et al. [A pathological report of three COVID-19 cases by minimally invasive autopsies], Zhonghua Bing Li Xue Za Zhi. 2020;49(0):E009.

8. Lefrancais E, Mallavia B, Zhuo H, Calfee CS, Looney MR. Maladaptive role of neutrophil extracellular traps in pathogen-induced lung injury. JCI Insight. 2018;3(3).

9. Xu J, Zhang X, Pelayo R, Monestier M, Ammollo CT, Semeraro F, et al. Extracellular histones are major mediators of death in sepsis. Nat Med. 2009;15(11):1318–21.

10. Connors JM, Levy JH. Thromboinflammation and the hypercoagulability of COVID-19. J Thromb Haemost. 2020.

11. Whyte CS, Morrow GB, Mitchell JL, Chowdary P, Mutch NJ. Fibrinolytic abnormalities in acute respiratory distress syndrome (ARDS) and versatility of thrombolytic drugs to treat COVID-19. J Thromb Haemost. 2020.

12. Varga Z, Flammer AJ, Steiger P, Haberecker M, Andermatt R, Zinkernagel AS, et al. Endothelial cell infection and endotheliitis in COVID-19. Lancet. 2020;395(10234):1417–8.

13. Chung M, Bernheim A, Mei X, Zhang N, Huang M, Zeng X, et al. CT Imaging Features of 2019 Novel Coronavirus (2019-nCoV). Radiology. 2020;295(1):202–7.

14. Buonsenso D, Pata D, Chiaretti A. COVID-19 outbreak: less stethoscope, more ultrasound. Lancet Respir Med. 2020;8(5):e27.

15. Borghesi A, Maroldi R. COVID-19 outbreak in Italy: experimental chest X-ray scoring system for quantifying and monitoring disease progression. Radiol Med. 2020.

16. Stertz S, Reichelt M, Spiegel M, Kuri T, Martinez-Sobrido L, Garcia-Sastre A, et al. The intracellular sites of early replication and budding of SARS-coronavirus. Virology. 2007;361(2):304–15.

17. Soeters PB, Wolfe RR, Shenkin A. Hypoalbuminemia: Pathogenesis and Clinical Significance. JPEN J Parenter Enteral Nutr. 2019;43(2):181–93.

18. Fulks M, Stout RL, Dolan VF. Albumin and all-cause mortality risk in insurance applicants. J Insur Med. 2010;42(1): 11–7.

19. Caironi P, Tognoni G, Masson S, Fumagalli R, Pesenti A, Romero M, et al. Albumin replacement in patients with severe sepsis or septic shock. N Engl J Med. 2014;370(15):1412–21.

20. Gorin AB, Stewart PA. Differential permeability of endothelial and epithelial barriers to albumin flux. J Appl Physiol Respir Environ Exerc Physiol. 1979;47(6):1315–24.

21. Yanagi S, Tsubouchi H, Miura A, Matsumoto N, Nakazato M. Breakdown of Epithelial Barrier Integrity and Overdrive Activation of Alveolar Epithelial Cells in the Pathogenesis of Acute Respiratory Distress Syndrome and Lung Fibrosis. Biomed Res Int. 2015;2015:573210.

22. Glas GJ, Van Der Sluijs KF, Schultz MJ, Hofstra JJ, Van Der Poll T, Levi M. Bronchoalveolar hemostasis in lung injury and acute respiratory distress syndrome. J Thromb Haemost. 2013;11(1):17–25.

23. Wu MA, Tsvirkun D, Bureau L, Boccon-Gibod I, Inglebert M, Duperray A, et al. Paroxysmal Permeability Disorders: Development of a Microfluidic Device to Assess Endothelial Barrier Function. Frontiers in medicine. 2019;6:89.

24. Radeva MY, Waschke J. Mind the gap: mechanisms regulating the endothelial barrier. Acta Physiol (Oxf). 2018;222(1).

25. Orsenigo F, Giampietro C, Ferrari A, Corada M, Galaup A, Sigismund S, et al. Phosphorylation of VE-cadherin is modulated by haemodynamic forces and contributes to the regulation of vascular permeability in vivo. Nat Commun. 2012;3:1208.

26. Aird WC. Phenotypic heterogeneity of the endothelium: II. Representative vascular beds. Circ Res. 2007; 100(2): 174–90.

27. Xie Z, Kuhns DB, Gu X, Otu HH, Libermann TA, Gallin JI, et al. Neutrophil activation in systemic capillary leak syndrome (Clarkson disease). J Cell Mol Med. 2019;23(8):5119–27.

28. Carsana L, Sonzogni A, Nasr A, Rossi R, Pellegrinelli A, Zerbi P, et al. Pulmonary post-mortem findings in a large series of COVID-19 cases from Northern Italy. The Lancet Infectious diseases. 2020; *In press*.

29. Shabbir W, Tzotzos S, Bedak M, Aufy M, Willam A, Kraihammer M, et al. Glycosylation-dependent activation of epithelial sodium channel by solnatide. Biochem Pharmacol. 2015;98(4):740–53.

30. Wolf T, Kann G, Becker S, Stephan C, Brodt HR, de Leuw P, et al. Severe Ebola virus disease with vascular leakage and multiorgan failure: treatment of a patient in intensive care. Lancet. 2015;385(9976):1428–35.Characteristics of the studied population.

